# Deciphering the Transcriptomic Landscape of Type 2 Diabetes: Insights from Bulk RNA Sequencing and Single-Cell Analysis

**DOI:** 10.1101/2024.11.06.24316740

**Authors:** A.A. Tkachenko, Z.N. Tonyan, Y.A. Nasykhova, Y.A. Barbitoff, I.N. Renev, M.M. Danilova, A.A. Mikhailova, O.B. Glavnova, S.V. Chepanov, S.A. Selkov, N.V. Golovkin, M.E. Vlasova, A.S. Glotov

## Abstract

Type 2 diabetes (T2D) is a chronic metabolic disorder marked by insulin resistance and relative insulin deficiency, affecting over 422 million people globally and projected to increase, making it a major public health concern. This condition is associated with severe complications such as retinopathy, nephropathy, cardiovascular diseases, and neuropathy, highlighting the need for a deeper understanding of its mechanisms to develop more effective prevention and treatment strategies. Transcriptomic analysis, particularly through RNA-seq, has provided valuable insights into the gene expression patterns in T2D, highlighting pathways involved in insulin signaling, metabolic regulation, and inflammation. This approach, including single-cell RNA sequencing, helps overcome the challenges of immune cell population heterogeneity, enabling the identification of distinct cell types and their specific roles in T2D progression. In this paper, we present a dataset of single-cell and bulk RNA sequencing results comparing expression patterns in blood cells between diabetes and healthy samples and describe some preliminary analysis of the dataset, highlighting differences in both gene expression and cell type composition and putting them in the context of existing research of T2D.

## 1. Introduction

Type 2 diabetes (T2D) is a chronic metabolic disorder characterized by insulin resistance and relative insulin deficiency. It is a significant public health concern worldwide, with an estimated prevalence of over 422 million individuals affected globally [1]. This number is projected to rise, making diabetes one of the leading causes of morbidity and mortality [2]. It is associated with numerous severe microvascular and macrovascular complications, including retinopathy, nephropathy, cardiovascular diseases, and neuropathy, which substantially impact patients’ quality of life and healthcare systems [3].

The global prevalence of T2D necessitates a deeper understanding of its underlying mechanisms to develop more effective prevention and treatment strategies. Recent advancements in transcriptomics and single-cell technologies have revolutionized our ability to investigate the cellular and molecular landscapes of T2D. Transcriptomic analysis provides insights into gene expression patterns in T2D, revealing pathways involved in insulin signaling, metabolic regulation, and inflammation. The importance of inflammation driven by immune cells in the pathogenesis of T2D has been repeatedly demonstrated [4, 5]. This makes research focused on the analysis of gene expression in whole blood cells especially relevant. Transcriptomic markers of T2D have been investigated in studies utilizing expression arrays and RNA-seq. While expression arrays allow for the analysis of a gene set limited by the design of microarrays, RNA-seq allows investigating the entire transcriptome. A study focused on the analysis of transcriptomic markers of T2D in whole blood using RNA-seq demonstrated differential expression of genes associated with inflammation, insulin resistance, and mitochondrial dysfunction [6]. However, the heterogeneity of immune cell populations poses significant challenges in understanding the disease’s complexity. Single-cell RNA sequencing (scRNA-seq) enables researchers to dissect this cellular diversity, allowing for the identification of distinct cell types and their specific contributions to T2DM progression.

By integrating transcriptomic data with single-cell analyses, we can elucidate the dynamic interactions between various cell types and their roles in T2DM. In this study, we aimed to analyze transcriptomic markers of T2D in whole blood cells and conduct single-cell sequencing on isolated PBMCs from T2D patients and healthy controls in order to advance our understanding of the processes involved in the pathogenesis of T2D and to highlight the significance of transcriptomic and single-cell research in T2D studies.

## 2. Results and discussion

Single-cell RNA-seq data was obtained from two diabetes patients and two healthy controls. After filtering based on quality control, 6974 cells remained. Uniform Manifold Approximation of Projection (UMAP) was used for visualization of PBMCs. Sample integration with Harmony did not show any pronounced batch effects that would differentiate cells from different individuals (Supplementary Figure S1). Clustering analysis using the Louvain algorithm predicted 15 clusters (Figure 1A). Cells were additionally annotated with Azimuth. Based on markers and Azimuth annotations cluster identities were predicted as follows: 0 - CD4+ T central-memory (TCM), 1 - CD4+ TCM and CD4+ naive cells, 2 - CD8+ effector-memory (TEM), 3 - natural killer (NK) cells, 4 - CD14+ monocytes, 5 - CD8+ naive T cells, 6 - CD14+ monocytes, 7 - intermediate B cells, 8 - naive B cells, 9 - CD16+ monocytes, 10 - mucosal-associated invariant T cells (MAIT), 11 - platelets, 12 - regulatory T cells, 13 - CD4+ TCM and CD14+ monocytes, 14 - plasmacytoid dendritic cells (pDCs).

**Figure 1.**
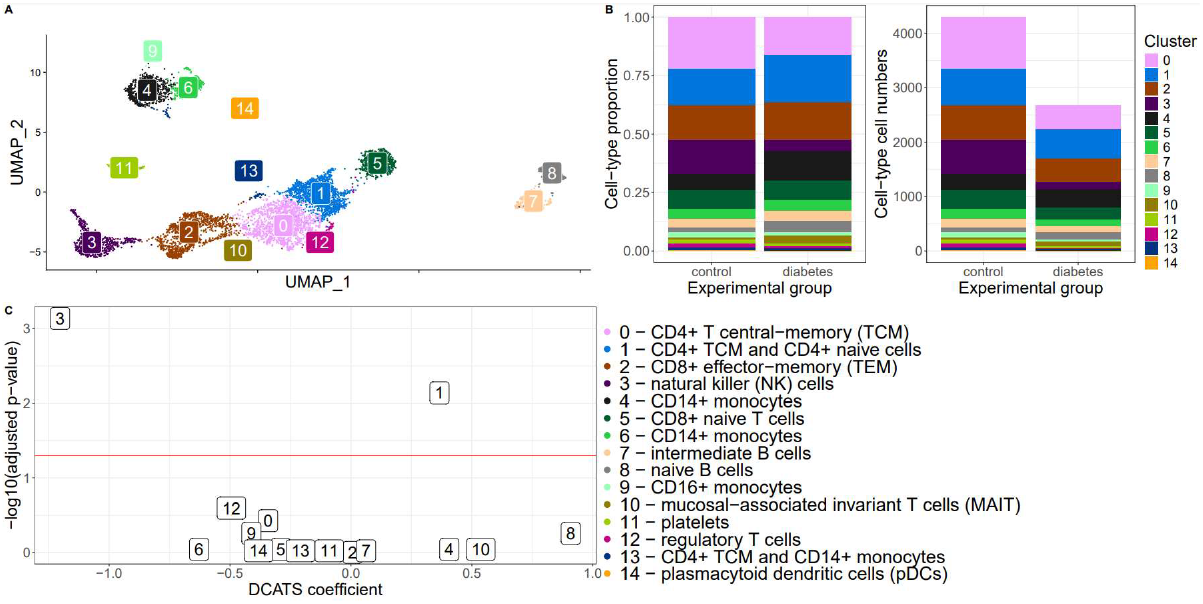
PBMC cell atlas and differences between cell type proportions between type 2 diabetes and healthy samples. A) UMAP projection for clusters of PBMC. Colors and numbers 0-14 correspond to respective clusters resulting from Louvain clustering done in Seurat. B) Proportions (left) and absolute amounts (right) of clusters after filtering. C) Results of differential composition analysis with DCATS. X axis shows value and direction of change in comparison of cell composition between diabetes samples and controls. Y axis is a negative log base 10 of p-value. Only clusters 3 and 1 pass the threshold of 0.05 (red line).

We compared whether diabetes and control samples differ in terms of cell type composition. Absolute counts for annotated cell types and percentages in diabetes and control samples are presented on Figure 1B. Healthy samples overall have more cells passing quality filters and some cell types’ proportions are noticeably different between sample groups. For a more rigorous comparison between groups of samples we calculated differential composition using DCATS for both Azimuth annotations of cells and our clustering. Directions of change and significance of differential composition are presented on Figure 1C for resulting clusters. A significant difference was observed for clusters 3 (NK cells) and 1 (CD4+ T central memory and naive cells) which means that NK cells were less prevalent in diabetes samples, and CD4+ T central memory and naive cells were more abundant in samples with T2D diagnosis.

In addition to single-cell RNA sequencing data on diabetes and healthy control we analyzed bulk RNA sequencing of 16 samples (8 diabetes and 8 controls, including two samples from single-cell data). For that read data was pseudoaligned to human reference with kallisto and differential expression (DE) was calculated with DESeq2. Volcano plot of differential expression results is presented on Figure 2A. Among 74163 genes with non-zero read counts, 146 genes pass the threshold of adjusted p-value < 0.05, with 71 genes increasing their expression in diabetes and 75 genes decreasing. PC biplot constructed with the top 500 most variable genes (Figure 2B) demonstrates that samples mostly mix and do not group according to their labels. Top 20 enriched GO terms are shown in Figure 2C.

**Figure 2.**
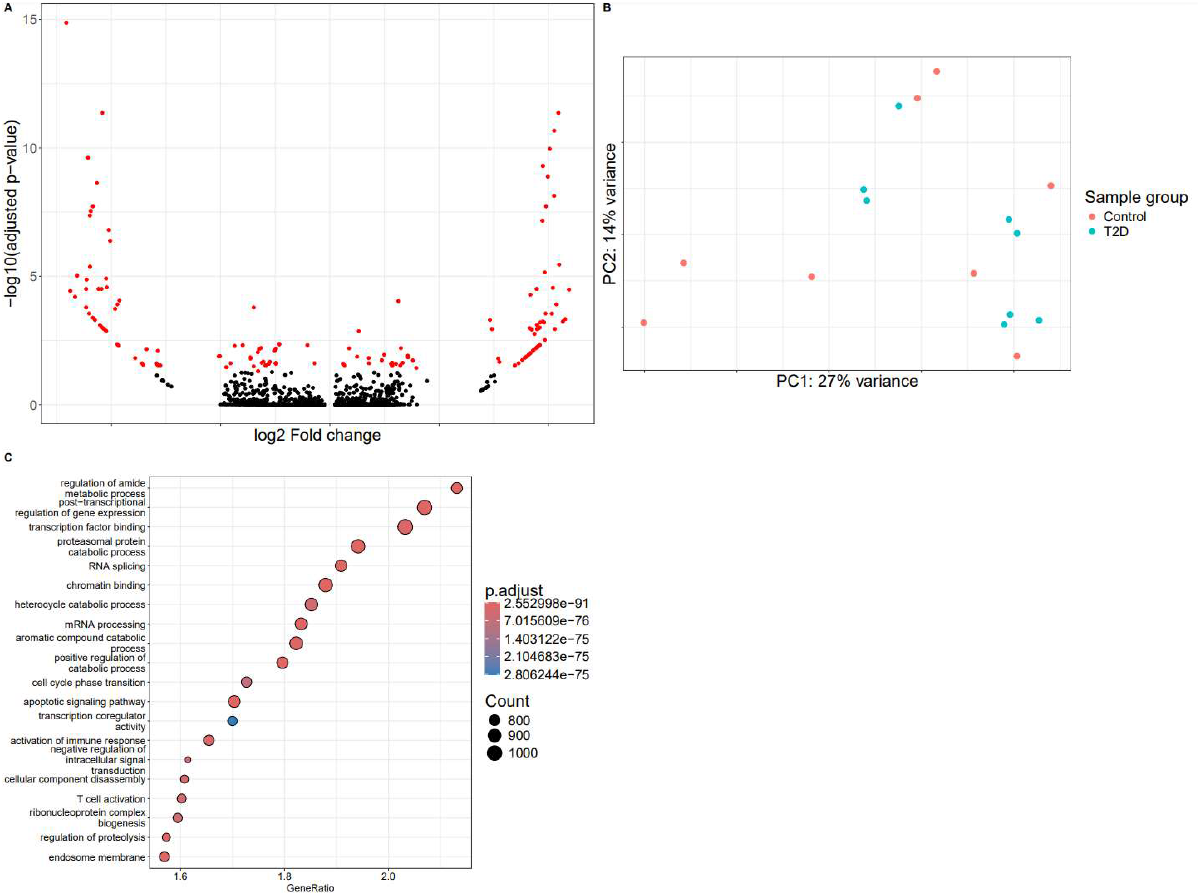
Comparison of bulk RNA sequencing between type 2 diabetes and healthy samples. A) Volcano plot of differential expression results for bulk RNA sequencing. Red dots correspond to genes with adjusted p-value < 0.05 in comparison between diabetes and control samples. B) PCA biplot showing distribution of samples based on top 500 most diverse genes. Samples are colored according to their group. C) Top 20 enriched GO terms based on differential expression results.

A similar study of PBMC single-cell RNA sequencing has been published recently [7]. One notable distinction of our data from results by Gu et al. is that we have observed a different trend of cell type composition, especially so in regard to NK cells which in our case are depleted in diabetes samples compared to healthy controls. This effect might be due to smaller sizes of sample groups in our comparison. As for the bulk RNA sequencing results, while some genes are differentially expressed between diabetes and controls it is worth mentioning that the majority of variance in the experiment is not explained by the disease status (Fig. 2B) and might be attributed to some other characteristics describing subjects.

## 3. Methods

### Study Cohorts and Participants

Peripheral blood samples were collected from 9 healthy volunteers and 9 unrelated individuals with T2D. T2D was diagnosed based on the World Health Organization criteria. The inclusion criteria for the control group were age over 30 years and no history of diabetes. Patients with type 1 diabetes, gestational diabetes, newly diagnosed diabetes mellitus (less than 1 year), acute or decompensated liver and kidney disease, autoimmune disorders, malignancies, and under 30 years of age were excluded from the study. The levels of fasting blood glucose (FBG), high-density lipoprotein (HDL), low-density lipoprotein (LDL), total cholesterol, and creatinine were measured in a fasting blood sample. Body height and body weight were measured, and the body mass index (BMI) was calculated in all the participants. The clinical and biochemical parameters of all the participants enrolled in the study are summarized in Table 1.

**Table 1.** Clinical characteristics of T2D patients and healthy controls.

| Characteristics | T2D (n = 9) | Controls (n = 9) | p-value |
| --- | --- | --- | --- |
| Age (years) | 66,3 | 56,6 | 0,35 |
| Female (n) | 5 | 5 | NA |
| Male (n) | 4 | 4 | NA |
| Family history (n) | 1 | 5 | NA |
| Fasting blood glucose* | 8,8 | 4,7 | 0,0096 |
| Total cholesterol (mmol/L) | 5,8 | 6,7 | 0,33 |
| HDL (mmol/L) | 1,5 | 1,6 | 0,75 |
| LDL (mmol/L) | 3 | 4,4 | 0,25 |
| Creatinine (mmol/L) | 0,1 | 0,08 | 0,14 |
| BMI (kg/m2)* | 32 | 24,5 | 0,0007 |
| WHR* | 1 | 0,76 | 0,01 |

\* p < 0.05 compared to the control group; NA - not applicable; Family history - family history of T2D in close relatives.

Single-cell sequencing was performed on PBMC samples obtained from two healthy controls and two patients with T2D. All individuals were female, with no family history of T2D. The average age was 36 years. Fasting glucose, total cholesterol, and creatinine levels did not differ significantly between the groups and remained within normal ranges. The body mass index (BMI) of healthy donors was normal, while the BMI in the T2D patient group was elevated (avg. 26.5 and 44, respectively).

Written informed consent for the research was obtained from all the recruited patients and healthy donors. The study was conducted in accordance with the Declaration of Helsinki, and approved by the Institutional Ethics Committee of D.O. Ott Institute of Obstetrics, Gynecology, and Reproductology (protocol #130 from 16 July 2020). This study was performed using large-scale research facilities #3076082 “Human Reproductive Health”.

### Blood sample collection

Whole blood samples were collected from the study subjects in anticoagulant Improvacuter K2EDTA 9 mL tubes (Guangzhou Improve Medical Instruments Co., Ltd., Guangzhou, China) after an overnight fasting period of 12 h. The blood samples (n=16) for the bulk RNA sequencing experiment were then immediately aliquoted in RNAse-free cryotubes (Fluidx Ltd., Cheshire, UK) and stored at −80 °C until use to prevent freeze– thaw cycles. The blood samples (n=4) for single-cell sequencing experiment were used immediately for single cell isolation. Samples from two participants (one of the subjects with T2D, the second was a healthy control) were analyzed using both approaches.

### Single cell isolation

PBMCs were isolated from whole blood samples within 2 hours by Ficoll-Paque medium density gradient centrifugation for 30 min at 500 g. The cells were resuspended in 1X phosphate-buffered saline containing 0.04% filtered bovine serum albumin to achieve the required cell concentration (700–1200 cells per 1 µL). The final cell concentration and viability were determined using a Countess II Automated Cell Counter (Thermo Fisher Scientific, Waltham, MA, USA).

### Total RNA isolation

Whole blood samples were retrieved from cryostorage and thawed at room temperature. The total RNA was isolated from 300 µL of whole blood using a TRIzol reagent (Invitrogen, Carlsbad, CA, USA) with chloroform, purificated with PureLink Total RNA Blood Kit (Invitrogen™, Carlsbad, CA, USA) following the manufacturer’s instructions, and eluted with 45 µL RNase-free water. Total RNA was tested for purity using the NanoDrop 2000 spectrophotometer (Thermo Fisher Scientific, Waltham, MA, USA) and quantified with Qubit™ RNA High Sensitivity Assay Kit and Qubit 2.0 Fluorometer (both Invitrogen™, Carlsbad, CA, USA).

### RNA-seq library preparation

Libraries were constructed from 150 ng of previously extracted total RNA using TruSeq Stranded Total RNA library preparation kit (Illumina, Inc., San Diego, California) according to the manufacturer’s protocol. Briefly, after ribosomal RNA depletion and RNA fragmentation, first and second strand cDNA were synthesized, and 3’ ends were adenylated, followed by adapter ligation and enrichment of DNA fragments. The library’s size distribution was analyzed with Agilent 2200 TapeStation and Agilent High Sensitivity D1000 ScreenTape (both Agilent Technologies, Inc., Waltham, MA, USA).

### Single cell RNA-seq library preparation

Cell suspension with a concentration of 1000 cells per µl was used to achieve a targeted cell recovery up to 9000 cells. Gel beads-in-emulsion (GEMs) containing uniquely barcoded gel beads and suspended cells were generated using a 10x Genomics Chromium Controller device followed by reverse transcription. Single-cell libraries were constructed using Chromium Next GEM Single Cell 3’ Reagent Kit v3.1 according to the manufacturer’s instruction. The main steps of library preparation included breaking GEMs, pooled cDNA fractions recovery, purification, and amplification, followed by enzymatic fragmentation, end-repair, A-tailing, adapter ligation, and final amplification. The quality of the single-cell libraries was assessed with Agilent 2200 TapeStation and Agilent High Sensitivity D1000 ScreenTape (both Agilent Technologies, Inc., Waltham, MA, USA).

### Sequencing

Indexed libraries were pooled at equimolar ratios. RNA-seq was conducted identically for both experiments on Illumina HiSeq 2500 System with 75-bp paired-end reads using HiSeq Rapid SBS Kit (all Illumina, Inc., San Diego, California).

### scRNA-seq data analysis

Read data was aligned to the GRCh38 human reference and quantified using CellRanger v. 8.0.0 [8]. Resulting data was analyzed with Seurat v. 5.1.0 [9]. Cells with more than 200 and less than 5000 features and less than 10% mitochondrial reads were kept for further analysis. A LogNormalize function with a scaling factor of 10,000 was applied to normalize the count matrices and identify variable features. First 2000 most variable features were used for PCA. Clusters were found using the Louvain algorithm with 0.5 resolution. The Harmony method was used to integrate datasets. Cells were assigned cell-type annotations with Azimuth v. 0.5 [10] using pbmcref as a reference. Differential cell composition between diabetes and control samples was calculated using DCATS v. 1.2.0 [11].

### Bulk RNA-seq data analysis

Bulk RNA sequencing data was pseudoaligned to the human GENCODE v46 reference with kallisto v.0.46.0 [12]. Differential expression was analyzed using DESeq2 package v. 1.44.0 [13]. Enrichment of GO terms in differential expression data was calculated using the GSEA method with clusterProfiler v. 4.12.6 [14].

## Data Availability

Single cell RNA sequencing raw data and CellRanger processing results are deposited in Gene Expression Omnibus under accession GSE280401. Bulk RNA sequencing raw data, counts and TPMs are deposited under GSE280402. Note that subject enumeration is continuous in these accessions and subject 2 and 4 are included in both types of experiments.

https://www.ncbi.nlm.nih.gov/geo/query/acc.cgi?acc=GSE280401

https://www.ncbi.nlm.nih.gov/geo/query/acc.cgi?acc=GSE280402

## Supplementary Materials

Figure S1: UMAP plot for distribution of cells among samples.

## Author Contributions

Conceptualization, G.A.S. and N.Y.A.; methodology, T.A.A. and T.Z.N.; software, T.A.A.; validation, B.Y.A. and N.Y.A.; formal analysis, T.A.A.; investigation, S.S.A., R.I.N., M.A.A., C.S.V., D.M.M., T.Z.N.; resources, G.N.V., V.M.E., G.O.B.; data curation, T.A.A., T.Z.N., G.O.B.; writing—original draft preparation, T.A.A. and T.Z.N.; writing—review and editing, B.Y.A., N.Y.A., and G.A.S.; visualization, T.A.A.; supervision, G.A.S. and N.Y.A.; project administration, G.A.S. and N.Y.A.; funding acquisition, G.A.S. All authors have read and agreed to the published version of the manuscript.

## Institutional Review Board Statement

The study was conducted in accordance with the Declaration of Helsinki, and approved by the Institutional Ethics Committee of D.O. Ott Institute of Obstetrics, Gynecology, and Reproductology (protocol #130 from 16 July 2020)

## Informed Consent Statement

Informed consent was obtained from all subjects involved in the study.

## Acknowledgments

We want to acknowledge Alexander Predeus for helpful discussion of the paper and we are deeply grateful to all the subjects of the study for their contribution.

