## Supplementary figures and images for "Deciphering the Transcriptomic Landscape of Type 2 Diabetes: Insights from Bulk RNA Sequencing and Single-Cell Analysis"

### Supplementary Figure S1

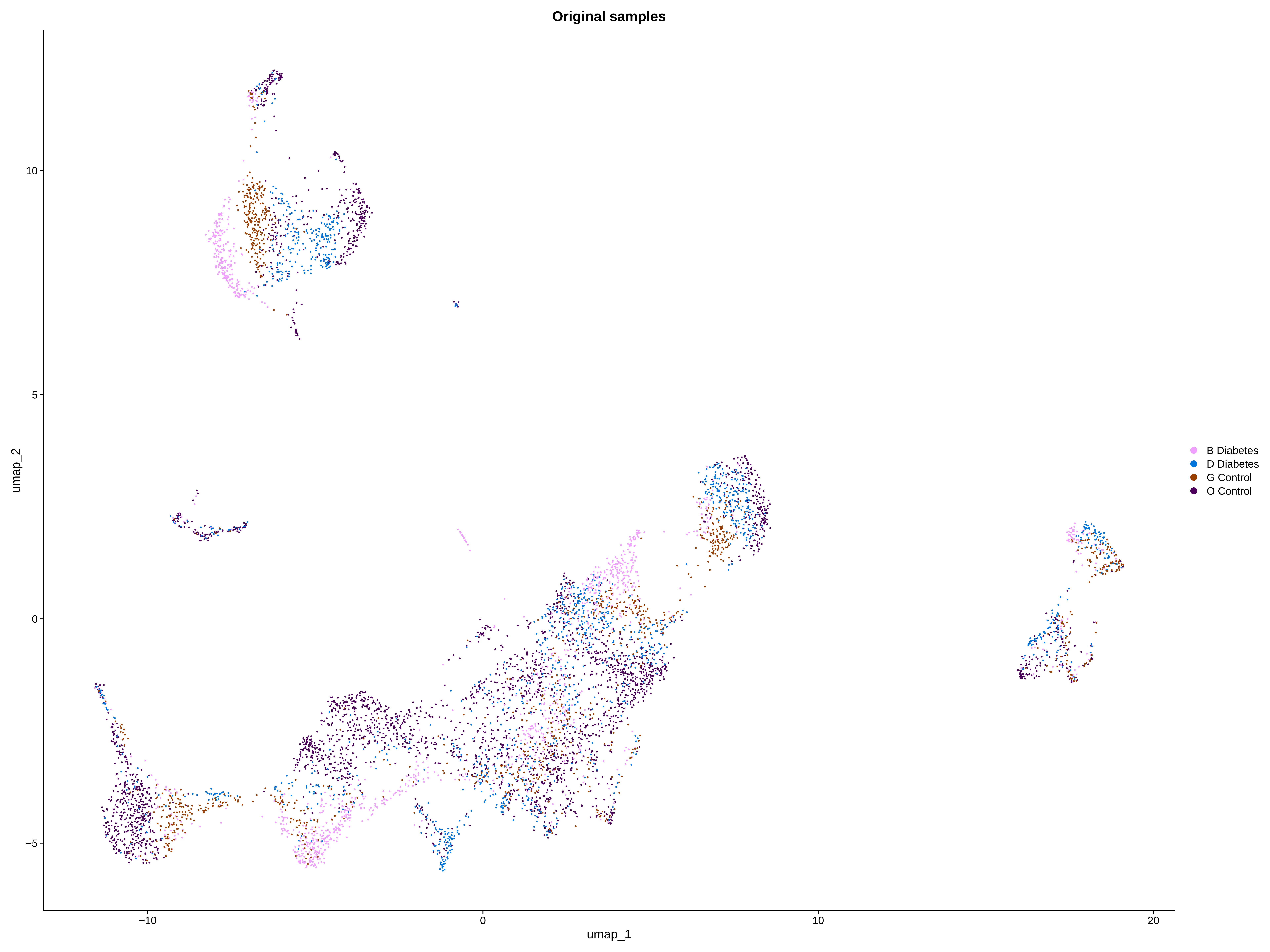
